# Racial-ethnic disparities in case fatality ratio narrowed after age standardization: A call for race-ethnicity-specific age distributions in State COVID-19 data

**DOI:** 10.1101/2020.10.01.20205377

**Authors:** Ishaan Pathak, Yoonjoung Choi, Dazhi Jiao, Diana Yeung, Li Liu

## Abstract

**Importance:** COVID-19 racial disparities have gained significant attention yet little is known about how age distributions obscure racial-ethnic disparities in COVID-19 case fatality ratios (CFR).

**Objective:** We filled this gap by assessing relevant data availability and quality across states, and in states with available data, investigating how racial-ethnic disparities in CFR changed after age adjustment.

**Design/Setting/Participants/Exposure:** We conducted a landscape analysis as of July 1st, 2020 and developed a grading system to assess COVID-19 case and death data by age and race in 50 states and DC. In states where age- and race-specific data were available, we applied direct age standardization to compare CFR across race-ethnicities. We developed an online dashboard to automatically and continuously update our results.

**Main Outcome and Measure:** Our main outcome was CFR (deaths per 100 confirmed cases). We examined CFR by age and race-ethnicities.

**Results:** We found substantial variations in disaggregating and reporting case and death data across states. Only three states, California, Illinois and Ohio, had sufficient age- and race-ethnicity-disaggregation to allow the investigation of racial-ethnic disparities in CFR while controlling for age. In total, we analyzed 391,991confirmed cases and 17,612 confirmed deaths. The crude CFRs varied from, e.g. 7.35% among Non-Hispanic (NH) White population to 1.39% among Hispanic population in Ohio. After age standardization, racial-ethnic differences in CFR narrowed, e.g. from 5.28% among NH White population to 3.79% among NH Asian population in Ohio, or an over one-fold difference. In addition, the ranking of race-ethnic-specific CFRs changed after age standardization. NH White population had the leading crude CFRs whereas NH Black and NH Asian population had the leading and second leading age-adjusted CFRs respectively in two of the three states. Hispanic population’s age-adjusted CFR were substantially higher than the crude. Sensitivity analysis did not change these results qualitatively.

**Conclusions and Relevance:** The availability and quality of age- and race-ethnic-specific COVID-19 case and death data varied greatly across states. Age distributions in confirmed cases obscured racial-ethnic disparities in COVID-19 CFR. Age standardization narrows racial-ethnic disparities and changes ranking. Public COVID-19 data availability, quality, and harmonization need improvement to address racial disparities in this pandemic.

**Key Points:** *Question:* What are the racial-ethnic disparities in COVID-19 case fatality ratios (CFR) across states after adjusting for age?

*Findings:* We conducted direct standardization among 391,991 COVID-19 cases and 17,612 deaths from California, Illinois and Ohio to compare age-adjusted CFR across race-ethnicities. The racial-ethnic disparities in CFR narrowed and the ranking changed after age standardization.

*Meaning:* Age distributions in confirmed cases obscured racial-ethnic disparities in COVID-19 CFR.

## Introduction

In the wake of the COVID-19 pandemic and Black Lives Matter protests, racial disparities in COVID-19 has received increased attention.^1,2,3,4^ The US COVID-19 response has been decentralized, with the federal government leaving responsibility to individual states. This has resulted in large variations in how states have collected, compiled and released COVID-19 data. As a result, publicly available data vary widely in content, format, and completeness, creating barriers for timely generation of evidence on racial disparities to inform policy and action. Still, existing data suggest the pandemic has disproportionately affected minority populations.

Among key indicators, racial disparities in case fatality ratio (CFR, defined as the number of COVID-19 confirmed deaths divided by the number of confirmed cases) is of particular significance as it represents the mortality risk of populations contracting SARS-CoV-2. CFR not only measures the severity of COVID-19 but also reflects differences in access and quality of care between populations.

Various factors, rooted in the systemic racism of US healthcare^5,6^ can drive racial disparities in CFR, including comorbidities^7,8,9^ and access to treatment.^10^ COVID-19 also has an age-dependent mortality pattern. Differences in the age distribution of cases by race may mask true racial disparities in CFR. The age distribution of the general US population varies by race, with Non-Hispanic (NH) White population being the oldest.^11^ Working-age NH White population are more likely to be able to work from home, while racial and ethnic minorities are more likely to be essential workers at the front line with low access to sick leaves^12^ or share households with someone who is^13^. These factors create variations in the age distribution of cases between race-ethnicities, with the older distribution of NH White cases potentially masking worse CFR in other races.

Existing evidence on how age confounds racial disparities in CFR is limited. One analysis compared age-specific COVID-19 mortality rates by race and found that after adjusting for race-specific age distributions, “Black people are dying from COVID at roughly the same rate as White people more than a decade older. Age-specific death rates for Hispanic/Latino people fall in between”.^14^ Another report further revealed that not only have Hispanic/Latino and NH Black populations been three times as likely to be infected as their NH White peers and twice as likely to die, but when comparing COVID-19 incidence rate among roughly the same age groups, this racial disparity widens further.^15^ However, neither of these studies examined age effects on racial disparities in CFR. Meanwhile, a multi-country study investigating the contribution of age to CFR variations concluded that the age distribution in confirmed cases often explains over two-thirds of CFR heterogeneity.^16^

In this study, we investigated racial disparities in CFR after controlling for age at the state level. We first assessed the availability and quality of data needed across states. ln states with appropriate age distributions of cases and deaths by race, we illustrated how racial disparities in CFR changed after age adjustment.

## Methods

### Landscape analysis of data availability and quality

A landscape analysis was conducted to assess data availability and quality from all 50 states and DC as of July 1, 2020. We reviewed the Department of Health and/or COVID-19 webpages from each state. To ensure we identified all relevant public information in any form (e.g., dashboards, reports, machine readable data files), we checked our findings against web searches of state COVID-19 data. We also cross-referenced against sources in the COVID Tracking Project’s racial data dashboard.^17^ We examined case and deaths data separately, and recorded the following: metrics (count vs. share of total); age-groups and race and/or ethnicity groups used for disaggregation; completeness (% of cases/deaths for which age-, race-ethnic, and age-race-ethnic-specific data were available vs. unknown); and downloadability.

We developed a grading system to assess availability and quality of these data. Five items for age- and race-ethnic-specific data were adopted respectively for a total score ranging from 0 to 10. The five items for age-specific data were whether: (1) any age-specific data existed; (2) for ages 50+, data were specific by <=10-year age increment; (3) the oldest, open-age group started at 80+; (4) data were disaggregated by 5-year age increments; and (5) unknown age was reported. The second and third items were the minimum data needed to control for different mortality risks in older ages. The fourth item was a convention in Demography. For race-specific data, the five items included whether: (1) any race-ethnic-specific data existed; (2) any ethnicity-specific data (i.e., Hispanic origin) existed; (3) Hispanic origin could be distinguished from other race-ethnicity combinations; (4) data were disaggregated by race and ethnicity combinations following federal guidelines;^18^ and (5) unknown race/ethnicity was reported. For example, if case data were reported across five racial groups (NH Asian, Hispanic, NH Black, NH White, and other), the state received a point for the third criterion. If the state additionally reported data by race across eight groups (Hispanic Asian, Hispanic Black, Hispanic White, Hispanic other, NH Asian, NH Black, NH White, and NH other), an additional score was assigned for the fourth criterion. Two co-authors independently assigned initial scores, then reviewed and reconciled the differences and arrived at the final scores.

### Adjusting age in race-ethnic-specific CFR by direct standardization

Age-and-race-ethnic-specific data on both cases and deaths were available in four states as of July 1, 2020: California^19^, Illinois^20^, Michigan^21^, and Ohio.^22^ However, data from Michigan did not distinguish Hispanic ethnicity from other races. For the other three states, we extracted age-and-race-ethnic-specific data on both cases and deaths manually.

To minimize idiosyncratic variations, we only included in the analysis a racial-ethnic group if 50+ cases were reported in each age-and-race-ethnic-specific group. As a result, we excluded American Indian & Alaskan Native, Hawaiian Native and Pacific Islander, and mixed race. The four race-ethnicity groups included were NH Black, NH White, Hispanic, and NH Asian. Ohio reported race and ethnicity based on federal guidelines^18^ and distinguishes Hispanic ethnicity for each race. To normalize this to the four race-ethnicity groups, we counted all Hispanic ethnicity as Hispanic race (regardless of stated race) and all the other cases (where ethnicity was NH or missing) as their stated race. California, Illinois, and Ohio applied CDC definitions of COVID-19 confirmed and probable case and death where relevant. Only Ohio made probable cases/deaths publicly available.

We used direct standardization to adjust for different age distributions among confirmed cases across race-ethnicities so that the CFRs were comparable.^23^ For each state, the total number of confirmed cases across all known race-ethnicities by age served as the standard population. Age-and-race-ethnic-specific CFRs were applied to this standard population to construct expected deaths within each age-and-race-ethnic group. Then the expected number of deaths were summed and divided by the total number of confirmed cases to construct age-standardized CFR as if all races had the same age distribution of confirmed cases.

We conducted two sensitivity analyses to better understand the impacts of biases due to incomplete reporting. We included both confirmed and probable cases/deaths using Ohio data (6% of total cases and 8.4% of total deaths). We also made a few assumptions about the non-trivial missingness in race-ethnicity. For example, in Illinois, race/ethnicity was self-identified at the time of testing and was missing for 50% of reported tests and 25% of confirmed cases.^24^ We treated “unknown race” as its own racial-ethnic group, divided unknown cases and deaths proportionately among existing racial-ethnic groups, and assumed certain extremes such as all unknown cases being NH White.

To illustrate how downloadable data could generate timely evidence during a pandemic, we also created a web-based dashboard application to automatically calculate age adjusted CFR with continuous updates. In States feasible, we wrote a Python script to scrape the COVID-19 race and ethnicity data reported daily, aggregated the data, and used the Python Data Analysis Library to calculate the age-standardized CFR.

## Results

### Data availability and quality

We found substantial variations by state in disaggregating and reporting case and death data by age and race-ethnicity. Figure 1 presents age-and-race-ethnic-specific COVID-19 case and death data availability and quality scores by state. Only California, Illinois, Michigan and Ohio reported age-and-race-ethnicity-specific data for both cases and deaths. Most states separately published age- and race-specific data.

**Figure 1.**
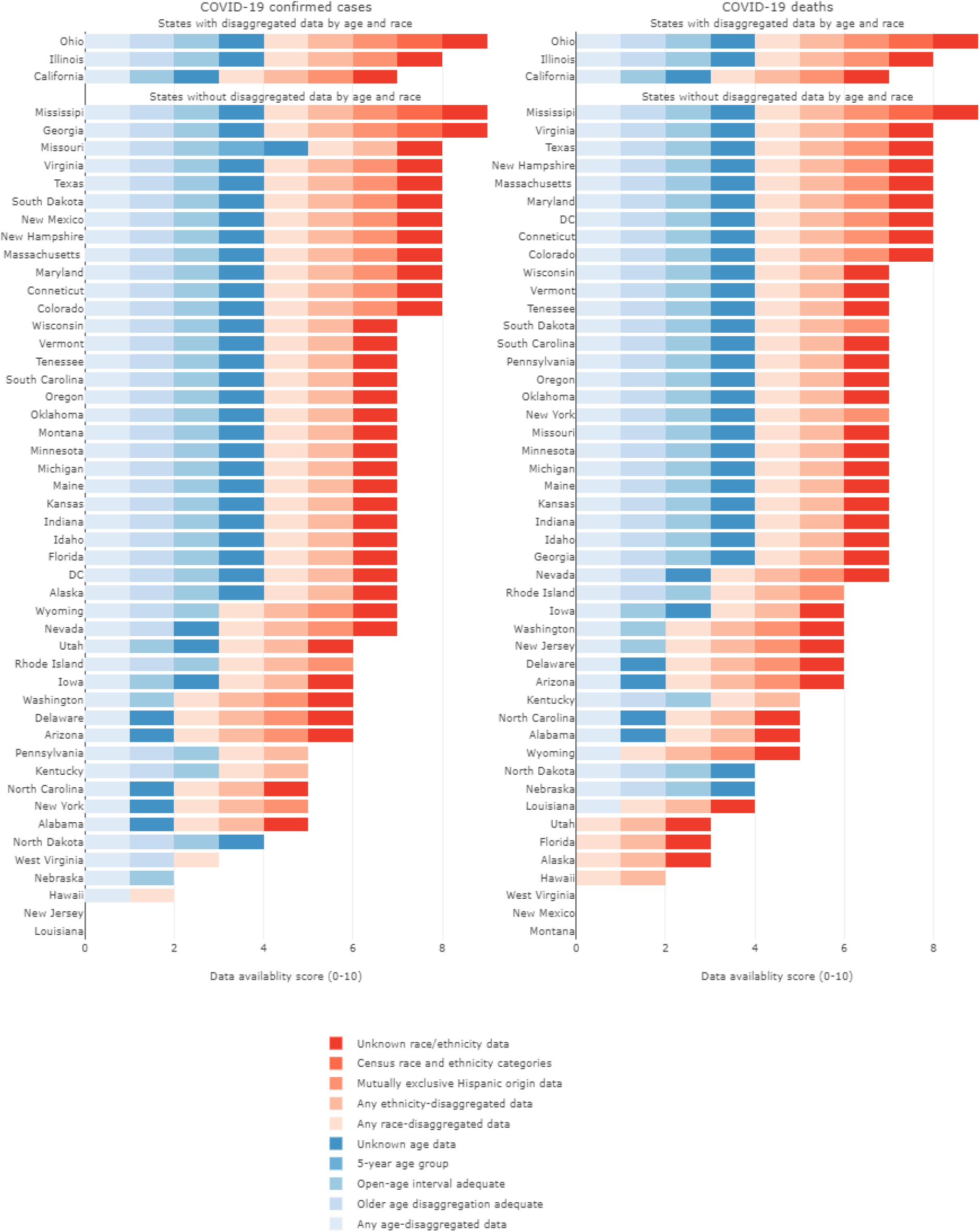
Availability and quality scores for age-specific and race-specific COVID-19 confirmed case and death data by states, US, 2020.

The average score was 6.4 (range: 0-9) and 6.0 (range: 0-9) for case and death data, respectively. Only 30 states had sufficient data to study the steep age-pattern of CFR in older populations (i.e., 10-year interval for age groups above 50+, and open-age interval starting at 80+). All states reported age-specific case data, but a few states, mostly with a small number of deaths, did not report age-specific deaths data. The choice of age groups varied widely. A majority of states used 10-year increments with others using 5-, 15- or 20-year ones. Even among states using the same increment, the exact age groups still varied (e.g., 20-29 vs 25-34). Some states did not use a consistent increment. The starting age of the open-age group also varied considerably, from 60+ in Hawaii to 100+ in Pennsylvania. A few states used inconsistent age-groups for cases and deaths, making it impossible to calculate age-specific CFR.

Nineteen states had sufficient data to study racial-ethnic disparities in CFR (i.e., data for Hispanic origin could be mutually exclusively distinguished from other race-ethnicity combinations). Many states reported race and ethnicity independent of each other, making it impossible to ascertain what proportions of cases/deaths within a given race were from Hispanic ethnicity. Four and five states did not report case and death data by race respectively. North Dakota and Nebraska did not publish race-specific data for cases or deaths at all. Few states reported race and ethnicity according to the federal guidelines, which reported and cross-tabulated both categories as in the censuses. Some states e.g. California also allowed the reporting of “Other”, an option not available in the census.

### Age-standardized racial disparities in CFR

Table 1 showed the descriptive statistics of our study data from California, Illinois and Ohio. In total, we analyzed 391,991confirmed cases and 17,612 confirmed deaths. While cases were spread evenly across ages, deaths concentrated among older age groups. Note the age grouping in California was different than the other two states and the much higher percentage of unknown race among cases than deaths.

**Table 1.** COVID-19 cases and deaths by age and race in California, Illinois and Ohio

|  | California |  | Illinois |  | Ohio |  |
| --- | --- | --- | --- | --- | --- | --- |
|  | Cases (%)<br>n=356,178 | Deaths (%)<br>n=7,283 | Cases (%)<br>n=156,963 | Deaths (%)<br>n=7,266 | Cases (%)<br>n=63,976 | Deaths (%)<br>n=2,813 |
| **Age Groups in California (years)** |  |  |  |  |  |  |
| 0-17 | 8.35 | 0.00 |  |  |  |  |
| 18-34 | 34.40 | 1.18 |  |  |  |  |
| 35-49 | 25.17 | 5.30 |  |  |  |  |
| 50-64 | 19.74 | 16.95 |  |  |  |  |
| 65-79 | 8.27 | 31.94 |  |  |  |  |
| 80+ | 3.94 | 44.64 |  |  |  |  |
| Unknown | 0.12 | 0.00 |  |  |  |  |
| **Age Groups in Illinois and Ohio (years)** |  |  |  |  |  |  |
| 0-19 |  |  | 8.24 | 0.06 | 7.98 | 0.06 |
| 20-29 |  |  | 17.42 | 0.37 | 19.7 | 0.35 |
| 30-39 |  |  | 16.45 | 0.74 | 16.85 | 0.74 |
| 40-49 |  |  | 17.25 | 1.55 | 14.62 | 1.55 |
| 50-59 |  |  | 16.40 | 6.28 | 14.70 | 6.28 |
| 60-69 |  |  | 11.40 | 14.05 | 11.24 | 14.05 |
| 70-79 |  |  | 6.33 | 24.65 | 7.20 | 24.65 |
| 80+ |  |  | 6.50 | 52.30 | 7.62 | 52.30 |
| Unknown |  |  | 0.01 | 0.00 | 0.09 | 0.00 |
| **Race and Ethnicity** |  |  |  |  |  |  |
| Latino/Hispanic | 35.34 | 43.13 | 31.6 | 20.6 | 7.07 | 2.24 |
| White | 11.40 | 30.44 | 22 | 44.5 | 44.42 | 74.23 |
| Asian | 3.77 | 13.21 | 2.79 | 4.79 | 2.86 | 1.14 |
| Black/AA | 2.80 | 8.62 | 16.80 | 27.7 | 26.81 | 19.69 |
| Multiracial | 0.47 | 0.56 |  |  |  |  |
| American Indian or Alaskan Native | 0.14 | 0.34 | 0.15 | 0.14 |  |  |
| Hawaiian Native or Pacific Islander | 0.39 | 0.48 | 0.25 | 0.11 |  |  |
| Other | 10.14 | 1.25 | 3.72 | 0.72 | 6.80 | 1.35 |
| Unknown/Missing | 35.56 | 1.96 | 22.7 | 1.41 | 12.05 | 1.35 |

The crude all-age CFR varied greatly by race and across states (Table 2). The largest racial-ethnic difference varied between 2.55 folds difference in California (7.27% among NH Asian population vs 2.57% among Hispanic population) to 5.29 folds difference in Ohio (7.35% among NH White population vs 1.39% among Hispanic population. However, the patterns of age-specific crude CFR were similar across the race-ethnicity groups, where the age-specific CFR increased substantially at 50+ (Figure 2). There were some distinct patterns across states. For example, Asians had the highest crude CFR at 80+ in California and Illinois, whereas NH Whites had the highest crude CFR at 60+ in Ohio.

**Table 2.** COVID-19 crude and adjusted CFRs by race in California, Illinois and Ohio

| Race | California |  | Illinois |  | Ohio |  |
| --- | --- | --- | --- | --- | --- | --- |
|  | Crude CFR | Adjusted CFR | Crude CFR | Adjusted CFR | Crude CFR | Adjusted CFR |
| Hispanic | 2.57 | 3.45 | 3.02 | 5.98 | 1.39 | 4.28 |
| NH Asian | 7.27 | 4.16 | 7.98 | 6.71 | 1.75 | 3.79 |
| NH Black | 6.56 | 4.34 | 7.67 | 6.95 | 3.23 | 4.70 |
| NH White | 5.63 | 3.01 | 9.49 | 5.53 | 7.35 | 5.28 |
| **Largest racial-ethnic disparity (fold)** | **2.55** | **1.44** | **3.14** | **1.26** | **5.29** | **1.39** |

**Figure 2.**
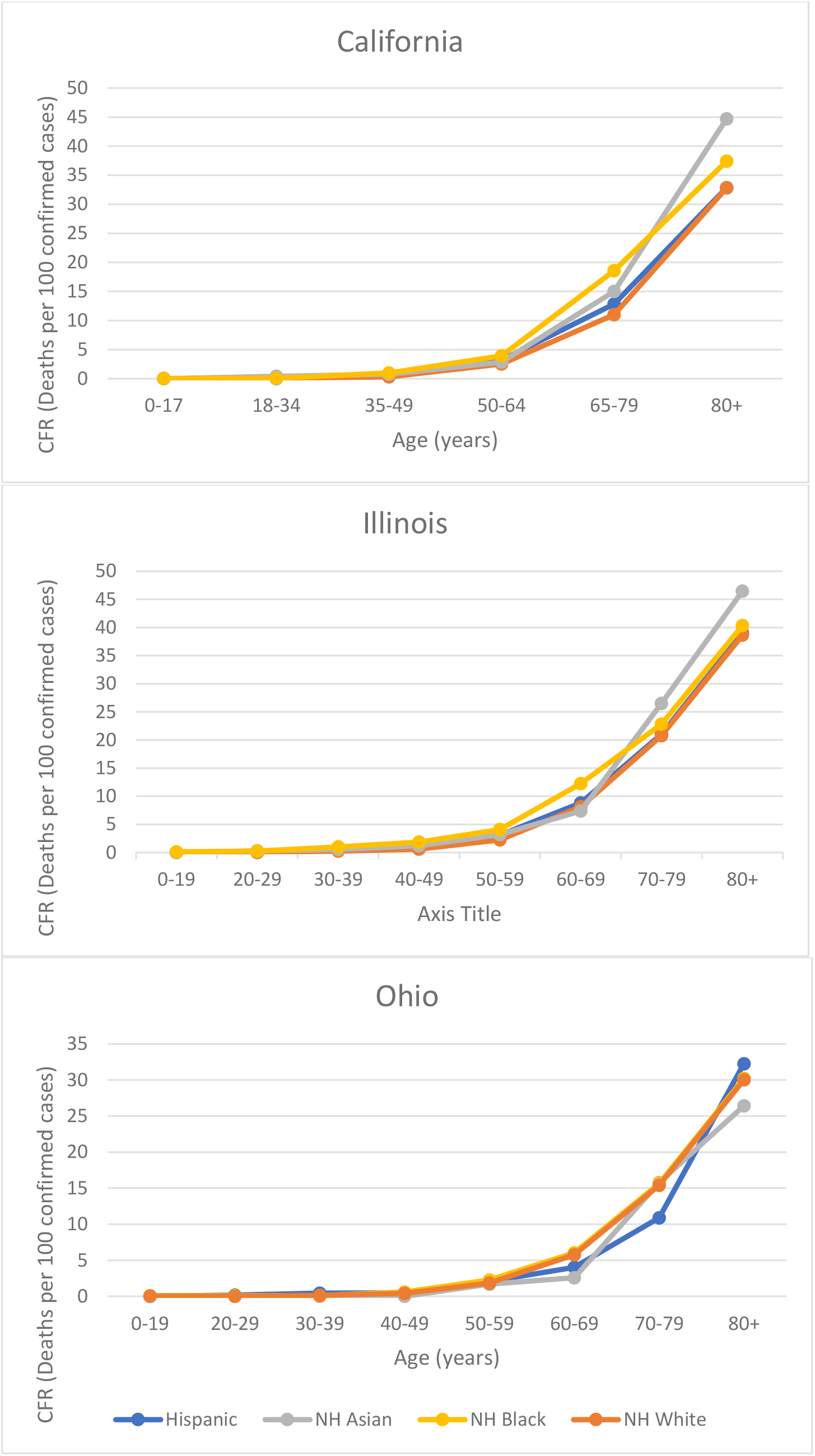
Crude age-specific CFR by race in California, Illinois and Ohio.

Racial-ethnic disparities in CFR narrowed after age-standardization. The largest racial-ethnic difference declined from 2.55 to 1.44 folds in California, from 3.14 to 1.26 folds in Illinois, and from 5.29 to 1.39 folds in Ohio (Table 2). The ranking of race-ethnic-specific CFR also shifted after age-standardization. In Ohio and Illinois, NH Whites had the highest crude CFR at 8.49% and 9.49%, respectively; In California, NH Whites had the third highest crude CFR at 5.63%. After age standardization, NH Whites’ CFR dropped to the lowest in California and Illinois at 3.08% and 5.52% respectively (Figure 3). NH Blacks had the second highest crude CFR in California (6.56%) and Ohio (3.23%), and the third highest in Illinois (7.66%). After age standardization, NH Black had the highest CFR in California (4.34%) and Illinois (6.95%), and the second highest in Ohio (4.70%). Hispanic populations had the lowest crude CFR in all three states and in California and Illinois, it was substantially lower than the second lowest CFR. After age adjustment the Hispanic CFR became the second lowest in all three states. NH Asians had relatively high crude CFR in California and Illinois. After age standardization, NH Asians’ CFR declined in both states but more than doubled in Ohio. The age-standardized CFRs were always higher than the crude CFRs among Hispanics, indicating the age distribution of Hispanic confirmed cases was younger than those of other races combined. The sensitivity analyses did not change the results qualitatively.

**Figure 3.**
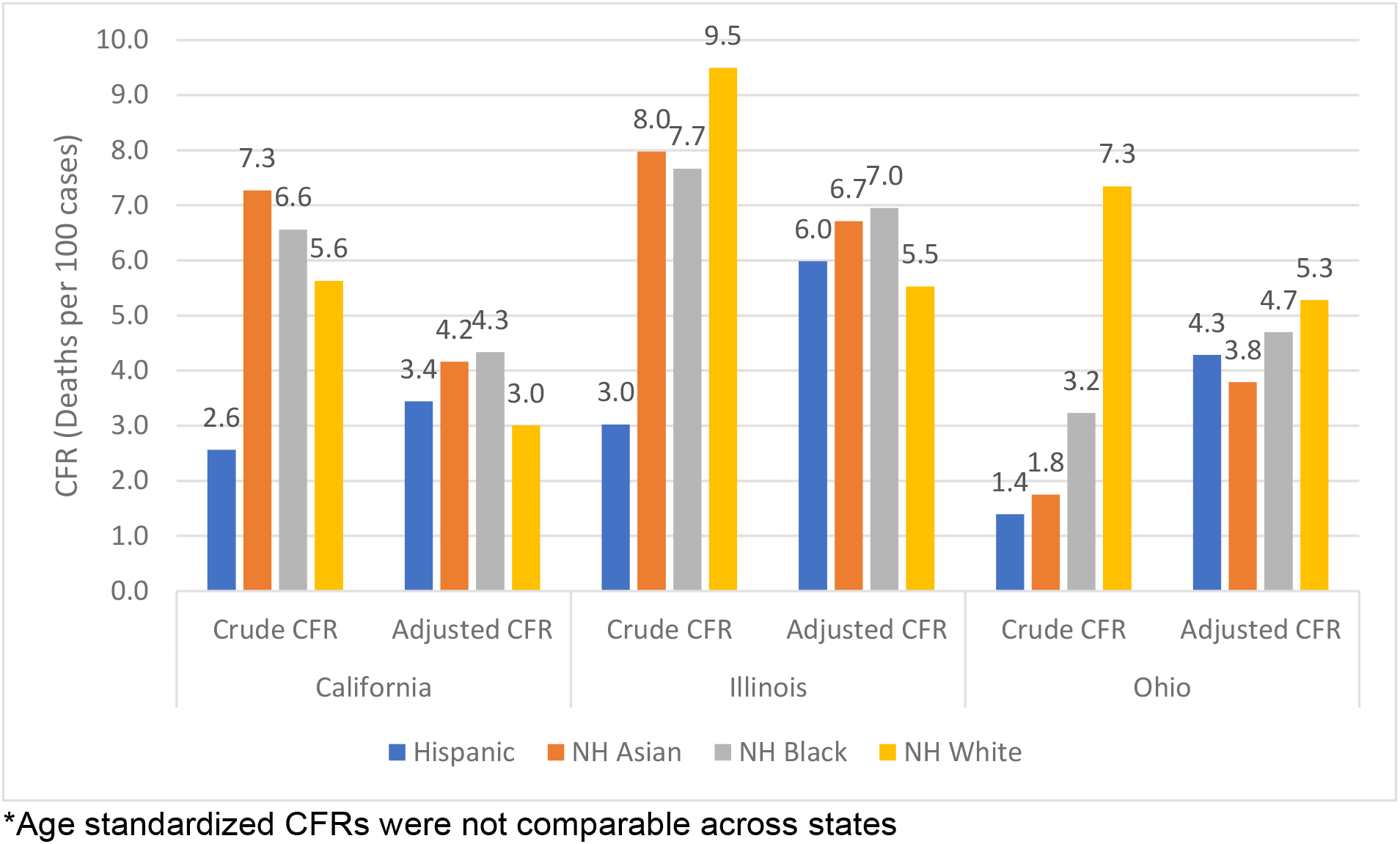
Crude and age standardized CFRs* by race in California, Illinois and Ohio.

Our online dashboard (https://adjusted-cfr.herokuapp.com/)^**Error! Bookmark not defined**.^ provides visualization of the crude and age-standardized CFR based on real-time data from California. The dashboard also presents age-and-race-ethnicity-specific cases, deaths, and expected number of deaths in each age group after age standardization for reproducibility. However, the data needed from Illinois and Ohio were embedded in dashboards or visualizations, which unfortunately prohibited automation of the age standardization and continuous updates.

## Discussion

In the US, more than five months after the first confirmed COVID-19 case,^25^ despite persistent racial disparities, only three states reported sufficient data to study racial-ethnic disparities in CFR controlling for age. Disaggregated data by either age or race were inconsistently available across states. Only thirty states had adequate data to study the steep age-pattern of CFR in old ages. Eighteen states had data to study CFR by race-ethnicity. It is critical to identify Hispanic ethnicity within racial categories as Hispanic populations have distinctive infection and mortality risks compared to other groups.

Age standardization in California, Illinois and Ohio revealed the obscuring effects of age distributions in confirmed cases on racial disparities in CFR. Racial disparities narrowed across all three states after age standardization and the ranking changed. The older age profile of NH White confirmed cases produced a higher crude CFR compared to other races. Upon age standardization, NH Whites had the lowest adjusted CFR in Illinois and California, and only slightly higher than NH Blacks in Ohio. Meanwhile the younger age profile of Hispanic cases led to much lower crude CFR than other races. Once age standardized, Hispanics had adjusted CFR comparable to others. NH Blacks’ CFR remained high or increased after age standardization. This aligned with previous evidence that Black communities have been disproportionately affected by COVID-19. NH Asians’ CFR remained high in California and Illinois after age standardization, a trend that has not been adequately documented. Data from more states is needed to understand the generalizability of these patterns.

Confirmed cases are only the tip of the iceberg of all COVID-19 infections. Multiple studies have documented the large number of asymptomatic and unreported COVID-19. In the US and other countries, COVID-19 deaths were less likely to be under-reported than cases. ^26,27,28^ Thus, the CFRs presented here could overestimate the mortality risk. However, it is unclear how this overestimation varied across age and racial-ethnic groups. While we cannot account for unreported cases, our sensitivity analysis using confirmed and probable cases in Ohio found no qualitative differences in the results. COVID-19 death is not a perfect measure either. Only 6%^29^ of deaths had COVID-19 as the only cause of death. The remainder had comorbidities as contributory causes and could be subject to misclassification of underlying cause of death. Conversely, deaths with COVID-19 as the true underlying cause could also be misclassified to other causes.^30^ Further investigation is needed to understand the interaction of these errors and how they distribute differentially across age and race-ethnicity.

Although age reporting among confirmed cases enjoyed high completeness, useful nuances, such as small increments in older ages and consistent age grouping, were often not available. More racial missingness was present among cases than deaths. If a majority of unknown race cases were from private facilities such as nursing homes where those on private insurance were overrepresented, these cases would likely be predominately NH Whites, leading to an overestimation of their CFR. Conversely, if inequitable access had prevented NH Black population from testing, their CFR’s could be overestimated.

Our analyses have two COVID-19 policy implications: public data availability and quality needs improvement and racial disparities in this pandemic must be addressed. On the data side, there is a need for higher resolution with improved completeness and intersectionality. Given the highly age-dependent nature of the COVID-19 pandemic, it is imperative for all states to publicly release age distribution of COVID-19 case and death data by race to gain a comprehensive understanding of racial disparities and their geographic variations in the US. There is also an urgent need to harmonize data across states to appropriately understand differences. Comparable data would also allow states to observe the effect policies in consideration have had on other states.

The landscape analysis revealed varying quantity, quality and format of state public COVID-19 data, causing challenges and delays in the generation of timely policy changes to curb the pandemic. Large age increments mask variations in CFR. Given the high variation in CFR between those in their 60s, 70s, and 80s, 80+ should be the minimum starting age for the open age group to uncover differences between late middle-aged and elderly populations. The format that public data was released in included different dashboards and/or downloadable files, making automatic data scraping difficult, rendering quick data analysis and fast evidence-based policy updates impossible.

As we improve on data reporting, we also need to actively address racial disparity and prioritize high-risk groups in healthcare and public health systems. Dowling et al^31^ emphasized that the response to COVID-19 cannot be “color-blind”. One specific policy to address racial-ethnic disparities could be to mandate race-ethnic reporting among cases as has been done for deaths. We did not include other races such as multiracial, Native Hawaiians or Pacific Islander, and Native Alaskans or American Indian in our analyses due to small numbers. However, some of these groups are also at heightened risk of COVID-19 mortality.

Despite narrowing after age standardization, racial-ethnic disparities were prevalent across all states considered. They indicated racial disparities in underlying comorbidities, healthcare access, and clinical treatment in the short term and chronic systemic racism in the long term. After accounting for age, NH Black population had consistently worse CFRs. Black communities need active consideration in the battle against COVID-19. For example, in New Orleans, clusters of COVID-19 cases in poor Black neighborhoods were missed by drive-through testing centers as many local residents did not own cars^32^. Now testing vans provide tests to these communities and other populations without cars.

Our analysis also found high CFRs among NH Asian population, particularly those at older age (70+). This phenomenon was reported to be more severe in urban areas including San Francisco and Chicago.^33^ Little is known on why these high CFRs are present. The NH Asian category encapsulates a huge range of cultures. More consideration must be applied to which among these groups are facing the brunt of the poor health outcomes. There has been documented stigma against certain Asian Americans due to COVID-19’s origins in China.^34^ This stigma could be affecting access to care. Other factors that may impact the high CFRs include low health literacy, low testing access and essential work positions among certain NH Asians.^35^

Publicly available, transparent and harmonized data practices coupled with policies and programs targeting high risk age groups among racial-ethnic minorities can increase our ability to better address racial disparities in COVID-19 and enhance life-saving responses to this dragging pandemic.

## Data Availability

All data used in this paper is publicly available. Primarily, all covid case and death data was taken from relevant state health department websites and is cited as such.

https://www.cdph.ca.gov/Programs/CID/DCDC/Pages/COVID-19/Race-Ethnicity.aspx

https://www.dph.illinois.gov/covid19/statistics

https://coronavirus.ohio.gov/wps/portal/gov/covid-19/dashboards/key-metrics/testing

https://rpubs.com/YJ_Choi/COVID19_US_DataAvailablityViz

## Acknowledgement

This research project is partially supported by the Johns Hopkins Population Center COVID-19 Lightning Pilot Award (R24HD042854).

## Conflict of Interest

None

## Authors’ contributions

Liu, Choi and Jiao conceptualized the study. Liu obtained partial funding for the study. Pathak carried out most of the analyses and wrote the first draft. Jiao conducted the automated data scraping and visualization. Yeung managed the project. All authors commented on subsequent drafts of the manuscript.

## Notes

### Competing Interest Statement

The authors have declared no competing interest.

